# A NEW PERSPECTIVE ON ISOTRETINOIN IN PREGNANCY: PREGNANCY OUTCOMES, EVALUATION OF COMPLEX PHENOTYPES, AND IMPORTANCE OF TERATOLOGICAL COUNSELLING

**DOI:** 10.1101/2023.06.02.23290862

**Authors:** Mustafa Tarık Alay, Aysel Kalaycı Yiğin, Mehmet Seven

## Abstract

**Background:** Teratogens are responsible for 5% of all known causes of congenital anomalies. Isotretinoin, a retinoic acid–derived agent, leads to congenital anomalies in 21-52% of cases when exposure occurs during pregnancy according to studies conducted before 2006. However, rates of congenital anomalies were much lower in later studies.

**Objectives:** To investigate the rates of congenital anomalies in isotretinoin exposure during pregnancy, isotretinoin exposure before pregnancy, and a control group unexposed to any teratogenic agents.

**Methods:** In this cohort study, we divided pregnant women admitted to our center between 2009 and 2020 into two groups: isotretinoin exposure during the pregnancy (n=77) and isotretinoin exposure before the pregnancy (n=75). We selected the control group from among the non-teratogen exposed pregnant women with a simple random sampling method. Obstetricians calculated the ages of all pregnancies via ultrasound (USG) (crown-rump diameter for the first trimester; biparietal diameter and femur length for the second trimester). After birth, a pediatric genetics specialist examined all babies.

**Results:** Among the isotretinoin exposure during the pregnancy, isotretinoin exposure before the pregnancy, and the control groups, there were statistically significant differences in live births (respectively, 64.3%, 88%, 93.3%), congenital anomalies (respectively, 28.6%, 6.1%, 1.4%), miscarriages (respectively, 13%, 2.7%, 4%), terminations (respectively, 32.5%, 9.3%, 2.7%), prematurities (11.9%, 16.7%, 2.9%) (respectively, p < 0.001, p<0.001, p=0.014, p<0.001). We detected novel phenotypical features in five patients.

**Conclusions:** Our study demonstrated that study design, long-term follow-up, teratological counseling, and implementing advanced molecular analysis in complex phenotypes with novel phenotypical features are beneficial for understanding the association of congenital anomalies with isotretinoin exposure. While evaluating congenital anomalies, we detected statistically significant differences between isotretinoin exposure vs control, but we did not detect any statistical differences between isotretinoin exposure before the pregnancy and controls. This conflict between our study and previous studies might be caused by no evident differentiation between isotretinoin exposure before the pregnancy and during the pregnancy and higher termination rates in previous studies.

## INTRODUCTION

Acne is a chronic inflammatory disease resulting from increased sebum production in pilosebaceous glands, keratinization disorder, inflammation^1^, and colonization of *Cutibacterium acnes*^2^. Isotretinoin, a retinoic acid-derived agent, affects all factors in acne pathogenesis and is the most effective treatment of severe recalcitrant acne^3^.

Isotretinoin is widely used in the pubertal period, adolescents, and women of childbearing age^4^. However, its teratogenic potential restricts its usage; therefore, isotretinoin in pregnancy is contraindicated^5^. Due to its sequestration in tissues and its long half-life, the cessation of isotretinoin at least one month before the onset of pregnancy is recommended, and reproductive-age women and their partners should be protected with contraceptive methods^6^. However, there is no evidence that isotretinoin exposure before pregnancy increases rates of congenital anomalies^7^.

Retinoic acid embryopathy (RAE), characterized by the cardiovascular, central nervous system, genitourinary system, thymic defects, microtia, and craniofacial defects, was first described by Lammer and colleagues in 1985^8^. Several novel phenotypic features in babies born to RA-exposed women in pregnancy were reported in later studies. However, these novel phenotypic features might stem from exposure to RA, genetic alterations, or incidentally^9–11^. Previously conducted genetic tests used for describing these novel phenotypic features had not been sufficient to discriminate the aforementioned reasons^8,10^. Therefore, more comprehensive molecular techniques need to be implemented^11,12^. Nowadays, whole exome sequencing (WES) is an advanced molecular technique developed for describing novel phenotypical features and illuminating the etiology of complex phenotypes^13,14^. Therefore, in this study, we performed WES to analyze novel phenotypic features in complex phenotypes in RA embryopathy patients.

In this study, we aimed to evaluate the association between congenital anomalies, miscarriages, prematurity, termination, and isotretinoin exposure in pregnancy, isotretinoin exposure before pregnancy, and a control group, and compare the results with each other. We also aimed to ascertain the role of teratological counseling in termination recommendations causing unwanted pregnancy losses without any medical indications.

## 2. MATERIALS and METHODS

### 2.1. Study Design and Patient Selection

This retrospective cohort study reviewed 12,678 pregnant women and child files admitted to our center between 2009 to 2020 for teratological counseling. The study group was selected from 152 of 177 retinoic acid-exposed pregnancies and divided into two categories: isotretinoin usage started before the pregnancy and cessation occurred before the corrected last menstrual period (LMP) as isotretinoin exposure before the pregnancy group (n=75) and those that continued their isotretinoin treatment after LMP as isotretinoin exposure during the pregnancy group (n=77). The control group (n=75) was selected from 2048 pregnant women by basic random sampling methods in non-isotretinoin exposure before or during the pregnancy, not taking C, D, X category medicines in their pregnancies, and not exposed to low-dose radiation (lower than 1 mGy) or other physical teratogenic agents.

Obstetricians calculated all gestational ages by USG based on corrected LMP. Calculations in the first trimester were implemented on crown-rump length (CRL), while second-trimester calculations were based on biparietal diameter and femur length. After the birth, a pediatric genetics specialist examined all live births. The study was conducted under the principles expressed in the Declaration of Helsinki. The institutional review board approved the study protocol with the decision number 06.01.2021/A-82(E-83045809-604.01.02-134912). Signed informed consent was obtained from all participants at the time of teratological consultation. The flowchart of the study is shown in Figure 1.

**Figure 1.**
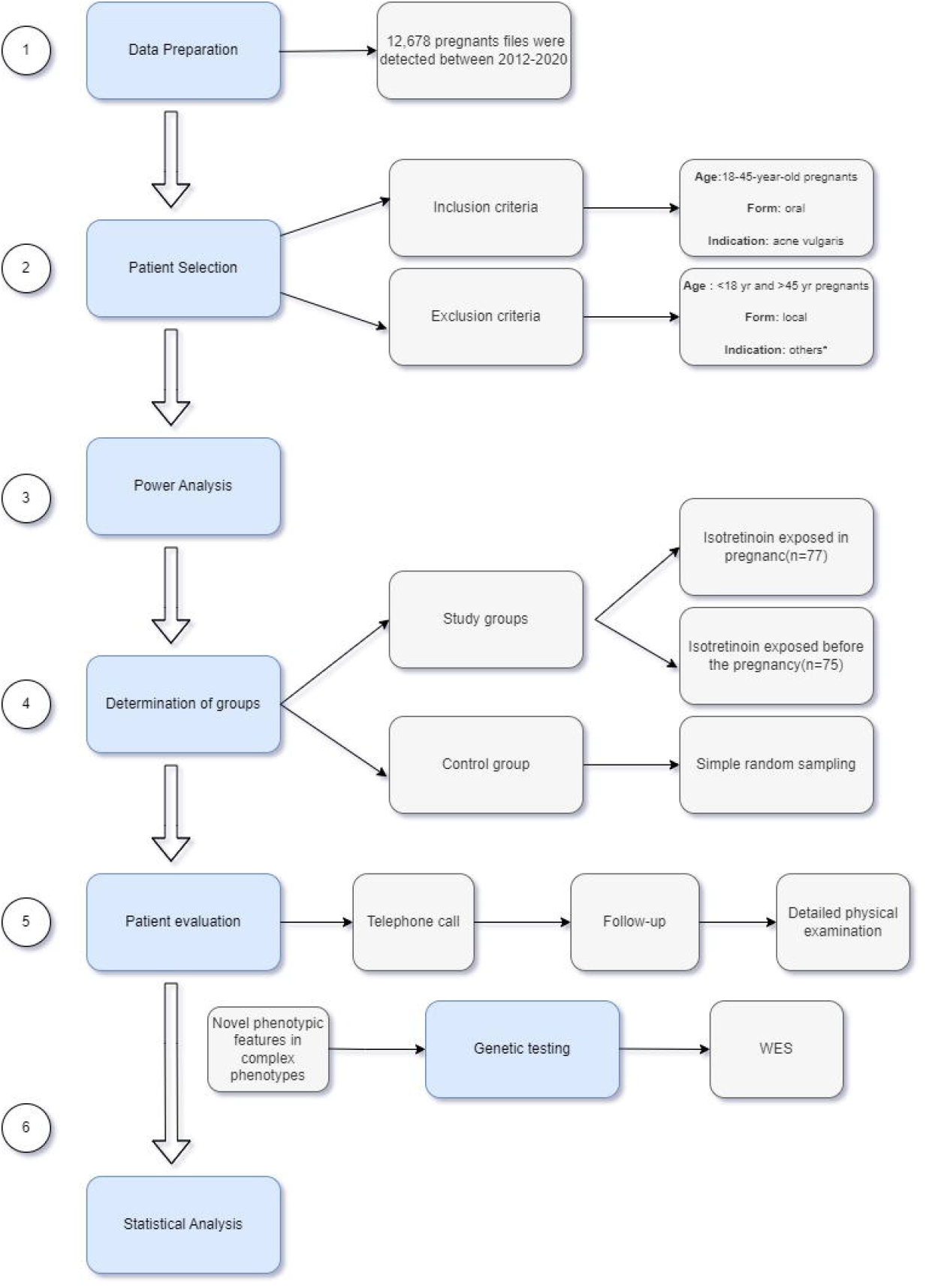
Flowchart of the study

### 2.2. Teratological Counseling and Follow-ups

Pregnant women who had a history of isotretinoin exposure in their anamnesis and were followed by gynecologists and obstetricians were referred to our center for teratological consultation. Potential teratogenic agents such as detailed drug use history, age, consanguineous marriage, chronic and/or hereditary disease history, smoking/alcohol/substance use, hyperthermia, and radiation exposure of the pregnant women who applied to our center were inquired about and recorded in teratological consultation files. Based on this information, a detailed teratological consultation report was prepared for the patients, and both the pregnant women and the medical specialists following the patients were informed in writing. After birth, babies were followed up by a pediatric genetics specialist, and their examination findings were recorded in their files.

### 2.3. Statistical Analysis

#### 2.3.1. Sample Size Calculation

Power analysis was implemented based on Zomerdijk et al. [15]. In this study, there were 4 anomalies in 51 babies whose mothers were exposed to isotretinoin in their pregnancies, and 9,041 anomalies in 208,108 babies whose mothers were not exposed to isotretinoin in their pregnancies. According to these values, the sync value was calculated as 0.1860465. The minimum sample size for each group was calculated as 35 in power analysis (95% CI and 5% alpha values).

#### 2.3.2 Statistical Analysis

SPSS 25 for Windows (IBM Corp. Released 2017. IBM SPSS Statistics for Windows, Version 25.0. Armonk, NY: IBM Corp.), GraphPad Prism version 8 for Windows (GraphPad Software, La Jolla California USA, www.graphpad.com), and Python 3.8.1 (pandas, matplotlib, and seaborn libraries) used for statistical analysis. All numerical variables (discrete or continuous) were tested for normality. If assumptions were met, means, and standard deviations were used; otherwise, medians and interquartile range were used. All categorical variables (nominal or ordinal) were expressed as numbers and percentages, and ordinal variables were sorted by their hierarchical placement in the tables. Both analytical (Kolmogorov-Smirnov test) and graphical methods (boxplot, Q-Q plot, detrended plot, histogram, and stem-leaf methods) were carried out for testing the normality assumption. Normality assumptions were made if skewness and kurtosis values were in the -1 to +1 range, and skewness and kurtosis indexes (skewness and kurtosis divided by standard error) were in the -2 to +2 range.

Student’s t-test and Analysis of Variance test (ANOVA) were conducted for normally distributed values. If ANOVA test outcomes were found to be statistically significant, then Tukey, Bonferroni, Sidak (equal variances assumed), and Duncan test (equal variances not assumed) were performed as a *post hoc* test. Mann-Whitney U test and Kruskal-Wallis H-test were carried out for non-normally distributed values. After the Kruskal-Wallis H test, the Dunn-Bonferroni test was performed as a *post hoc* test.

Chi-square and Fisher Freeman Halton test (if any expected values were smaller than 5) were performed for categorical variables analysis. Cramer V was conducted for a measure of association between two nominal variables. Confidence interval (CI) was determined as 95% for all statistical analyses, (α) and (1-β) values were accepted as 0.05 and 0.80, respectively. P values less than 0.05 (p<0.05) were accepted as statistically significant.

## 3. RESULTS

### 3.1. Descriptive Statistics

The mean ages of the isotretinoin exposure before the pregnancy group, isotretinoin exposure in the pregnancy group, and the control groups were 27.533 ± 0.594 (18-39), 27.481 ± 4.433 (19-38), and 27.467 ± 0.591 (18-37) years, respectively. Descriptive statistics are shown in Table 1, and the cumulative dose distribution of isotretinoin exposure in pregnancy is in Figure 2.

**Table 1.** Descriptive variables

| **Numerical variables** | **Isotretinoin exposure before the pregnancy**<br>**n=75**<br>**(mean ± SD)** | **Isotretinoin exposure during pregnancy**<br>**n=77**<br>**(mean ± SD)** | **Control group**<br>**n=75**<br>**(mean ± SD)** | **P values** |
| --- | --- | --- | --- | --- |
| Age | 27,533 ± 5,147 | 27,481 ± 4,433 | 27,467 ± 5,116 | .995<sup>1</sup> |
| **Categorical variables** | **Isotretinoin exposure before the pregnancy**<br>**n=75**<br>**(n,%)** | **Isotretinoin exposure during pregnancy**<br>**n=77**<br>**(n,%)** | **Control group**<br>**n=75**<br>**(n,%)** | **P values** |
| **Consanguinity** | 6 (7.9) | 9 (11.7) | 5 (6.3) | .526<sup>2</sup> |
| **Contraceptive method** | 58 (77.3) | 51 (66.2) | 35 (46.7) | <.001<sup>2</sup> |
| **Assisted reproductive techniques** | - | - | 5(6.7) | .007<sup>3</sup> |
| **Menstrual cycle** |  |  |  |  |
| Regular | 67 (90.3) | 64 (83.1) | 74(98.7) | .002<sup>3</sup> |
| Irregular | 8 (10.7) | 13 (16.9) | 1(1.3) |  |
| **Folic acid** | 50 (66.7) | 29(37.7) | 38 (50.7) | .002<sup>2</sup> |
| **Multivitamin** | 2(2.7) | 8(10.4) | 4 (5.3) | .148<sup>2</sup> |

1. One-way ANOVA test
2. Pearson chi-square test
3. Fisher exact test

**Figure 2.**
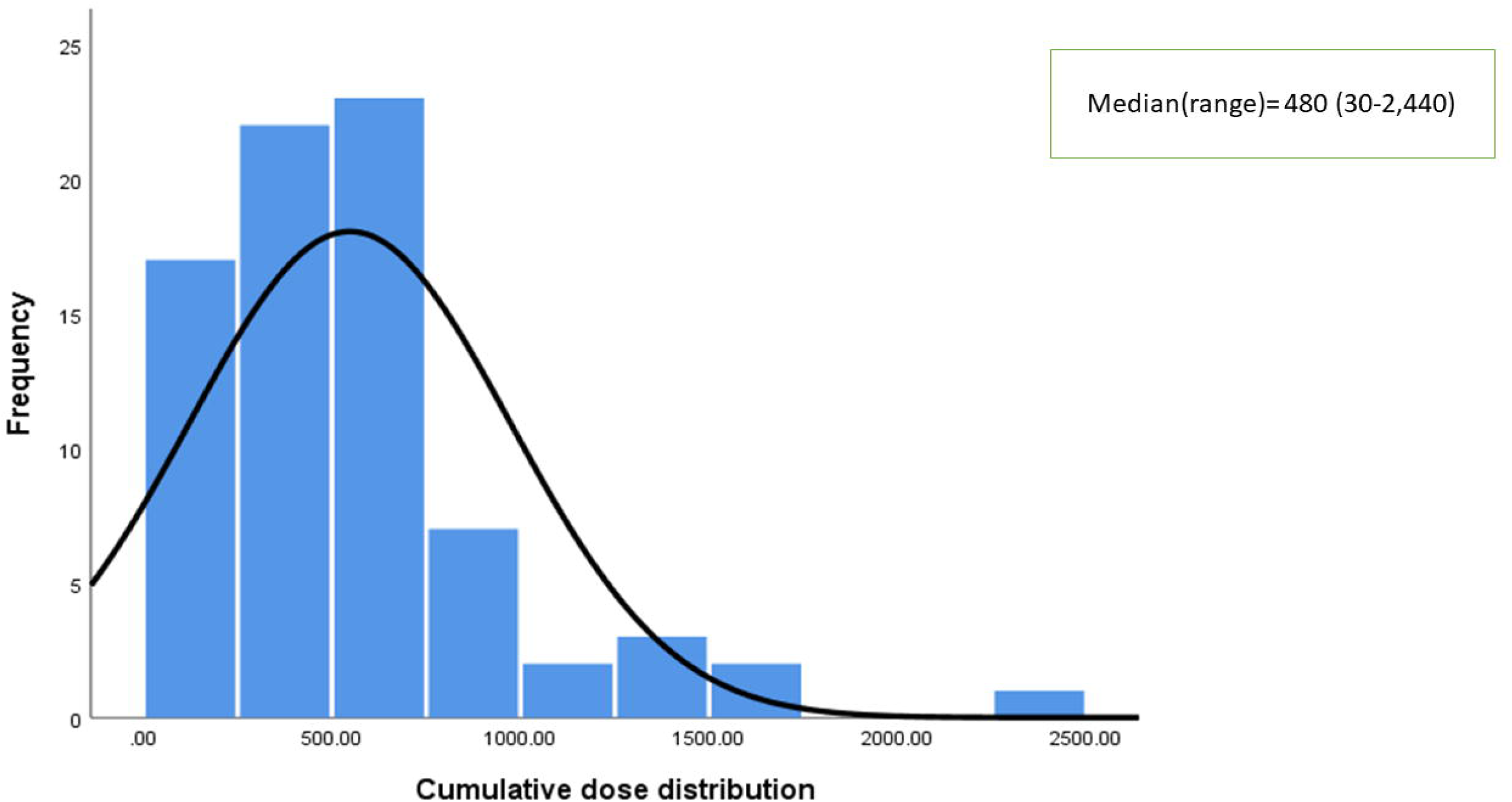
Cumulative dose distribution of isotretinoin exposure in pregnant women

**Figure 3.**
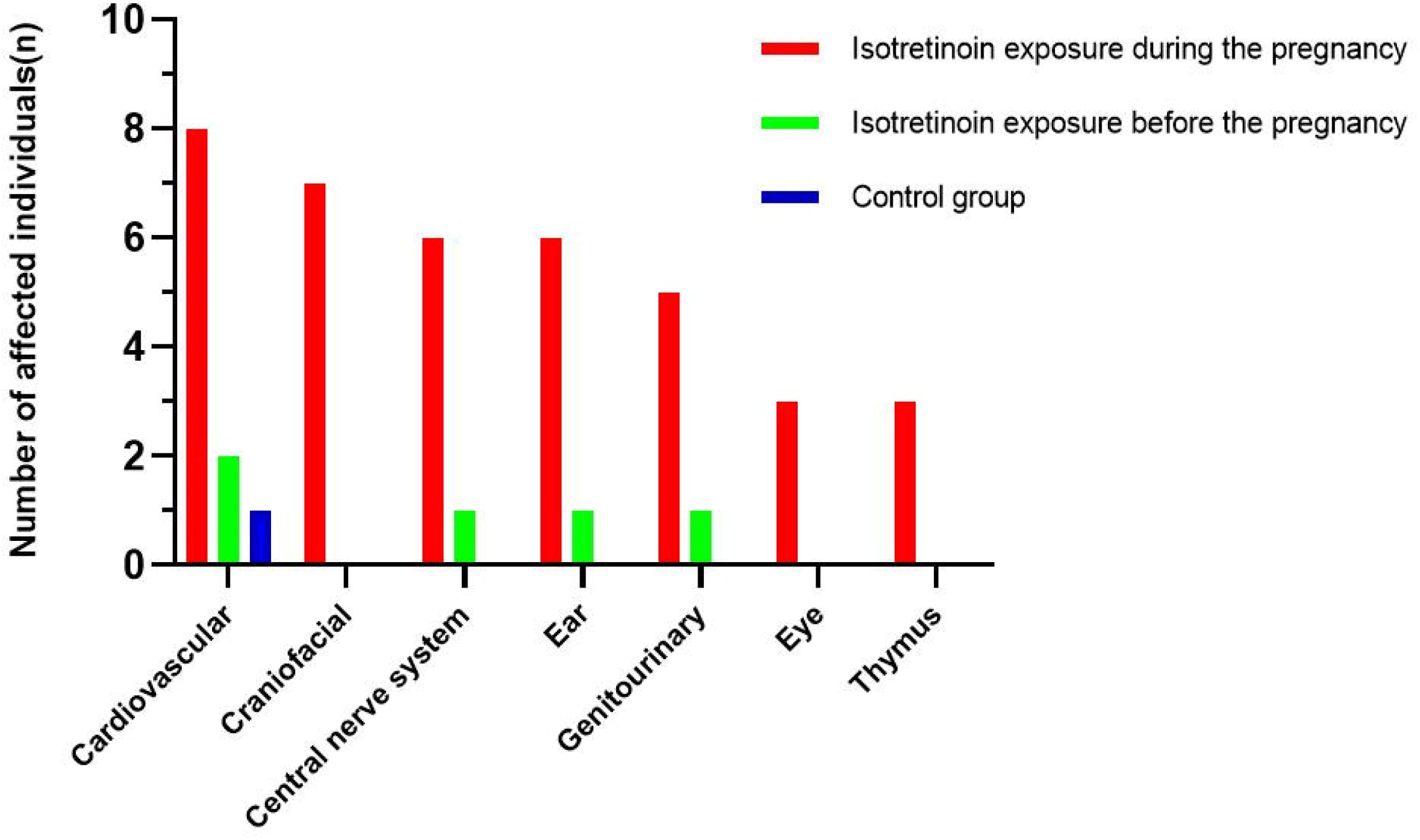
Distribution of congenital anomalies by individuals.

### 3.2. Pregnancy outcomes and evaluation of newborn fetal examination

Pregnancy outcomes are indicated in Table 2. Here, 42 of 77 (54.5%) of those with isotretinoin exposure during pregnancy, 66 of 75 (88%) of those with isotretinoin exposure before the pregnancy, and 70 out of 75 (93.3%) controls gave livebirth. When the groups were evaluated for live births, there was a statistically significant difference detected among the groups, and we found a moderate association between live birth and isotretinoin exposure period *X2* (2, N= 227) = 39.851, p<0.001, Cramer’s V= 0.419. The difference among groups stemmed from isotretinoin exposure during pregnancy.

**Table 2.**
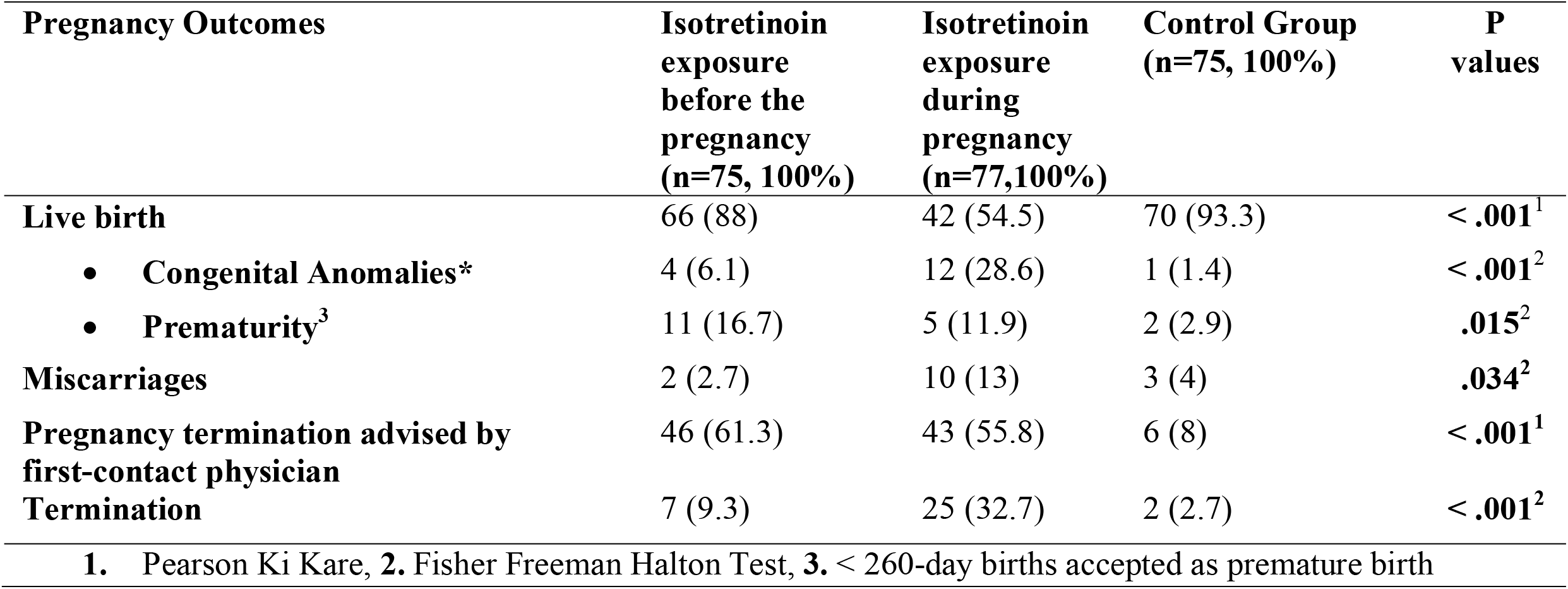
Pregnancy outcomes and neonatal characteristics

Congenital anomalies were found in 12 out of 42 (28.6%) live births in the isotretinoin exposure during the pregnancy group, 4 out of 66 (6.1%) live births in the isotretinoin exposure before the pregnancy group, and 1 out of 70 (1.4%) live births in the control group. When the groups were compared for congenital anomalies in live births, there was a statistically significant difference detected among the groups *X2* (2, N= 178) = 20.095, p<0.001, Cramer’s V= 0.366. This difference stemmed from isotretinoin exposure during pregnancy. The result indicated a moderate association between congenital anomalies and the isotretinoin exposure period. Congenital anomalies are shown in Table 1.

Comparing among the groups for prematurity, termination, and miscarriages, there was a statistically significant difference found between the groups (respectively, *X*^*2*^ (2, N= 178) = 7.810, p = 0.015, Cramer’s V = 0. 203, *X2* (2, N= 227) = 29.298, p<0.001, Cramer’s V= 0.359., *X2* (2, N= 227) = 6,801, p=0.034, Cramer’s V = 0.185).

### 3.3. Evaluating genetic variants in novel phenotypical features

We evaluated novel phenotypical features in babies born from isotretinoin-exposed women, most of them previously not reported in the literature (PubMed, Scopus, Web of Science). These are sixth nerve palsy, non-febrile seizures, stereotypical movements, asymmetric crying faces (very rare), and bilateral undescended testicles. We implemented WES analysis in 5 out of 7 patients characterized with complex phenotypes. We did not conduct WES analysis of the remaining two cases because they died in the early months of their lives. Detected genetic variants that might be associated with novel clinical features are shown in Table 3. All patients on whom WES was conducted had previously undergone G-banding karyotype analysis and microarray test results were normal.

**Table 3.** Results of WES in RA embryopathy patients with novel phenotypical features

| **Patients** | **Variants<sup>1</sup>** | **Prediction** | **Clinical signs** | **OMIM** |
| --- | --- | --- | --- | --- |
| **P-1** | TRPC6(NM_004621.6): c.1156C>T | LP | Focal segmental glomerulosclerosis | #603965 |
| **P-1** | SCN8A<sup>2</sup>(NM_001330260.2): c.1421C>G (p.Pro474Arg) | VUS-LP | Benign epileptic seizures | #617080 |
| **P-2** | LP/P and/or VUS variants associated with clinical features not detected | - | - | - |
| **P-3** | SCN1A<sup>2</sup>(NM_006920.6):c.2854G>T (p.Ala952Ser) | LP | Seizures | #604403 |
| **P-4** | LP/P and/or VUS variants associated with clinical features not detected | - | - | - |
| **P-5** | FV(NM_000130.5): c.1001G>C(p.Arg334Thr) | Pathogenic | Prone to thrombosis | #227400 |
| **P-5** | SAG(NM_000541.5): c.1103-2A>C | Pathogenic | Visual problems | #613758 |
| **P-5** | ALAS2(NM_000032.5): c.1435C>T(p.Arg479Trp) | VUS-LP | Anemia | #301300 |
| **P-5** | CCBE1(NM_133459.4):c.845G>A (p.Arg282Gln) | VUS-LP | Undescended testis, ectopic kidney, VSD | #235510 |
| 1. All variants were heterozygous state. |  |  |  |  |
| 2. Could be associated with novel phenotypic features |  |  |  |  |

## 4. DISCUSSION

This is the first cohort study that compared pregnancy outcomes, termination rates, and termination recommendations in isotretinoin exposure during pregnancy and isotretinoin exposure before the pregnancy with controls. Our study revealed some novel phenotypical features that have not been reported previously in babies born from isotretinoin-exposed pregnancies and tried to clarify the etiopathogenesis of the new findings by performing WES in these patients. Our study is also unique in this respect.

This study demonstrates that the congenital anomaly rates in babies born of isotretinoin-exposed pregnant women are statistically significantly increased compared to rates in babies born from women exposed to isotretinoin before the pregnancies and babies born in the control group. There are few studies available investigating isotretinoin exposure in pregnancy and congenital anomalies^15^. High congenital anomaly rates in isotretinoin exposure in pregnancy were based on studies before 2006, with rates of 21-52% reported^8,16,17^.However, lower rates of anomalies (0-17%) were found in recent studies^5,18–26^ [Table 4]. Among the last five studies^18,19,21,22,24^, congenital anomalies were not detected in two^19,24^, while congenital anomalies were found in one at a rate of 4%^21^. In our study, the rate of congenital anomalies detected in isotretinoin exposure during pregnancy was 28.6%, which is similar to the results of the studies before 2006.

**Table 4.**
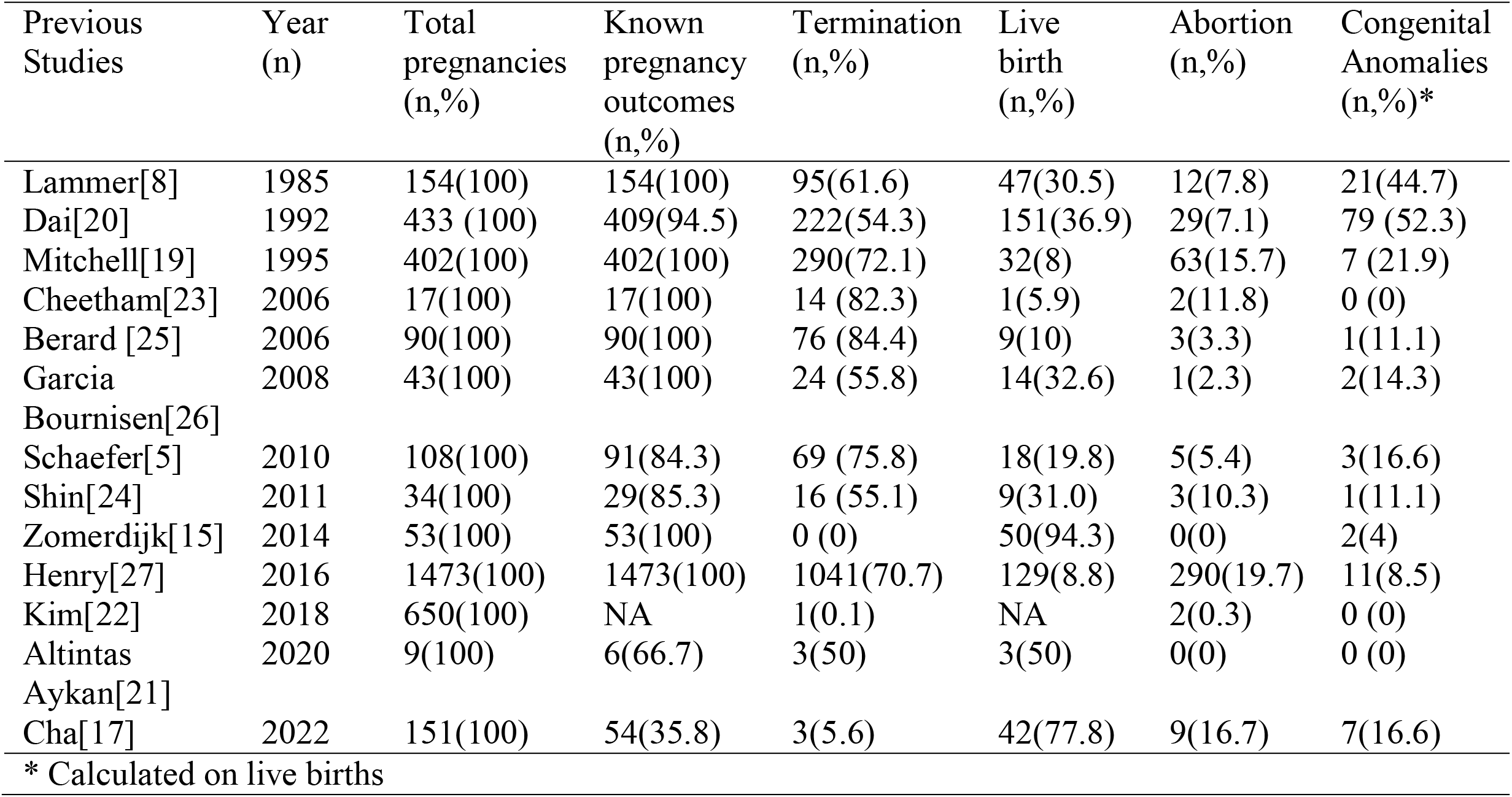
Previous studies on isotretinoin exposure in pregnancy

Although an increase in congenital anomaly rates was observed in the children of pregnant women exposed to isotretinoin before the pregnancy compared to the control group, this rate was not statistically significant. In our study, the rates of congenital anomalies in isotretinoin exposure before the pregnancies were 6.1%, similar to studies conducted after 2006. Detecting lower anomaly rates in studies carried out after 2006 than in past studies might stem from study groups not discriminating before the pregnancies and during pregnancy, high termination rates, and most of the pregnancies being terminated in early part of the pregnancy. Other causes include isotretinoin exposure in pregnancies that occurred during the organogenesis period, regular follow-ups, physicians not wanting a risk, some anomalies in pregnancies resulting in miscarriages and these miscarriages were not examined for congenital anomalies, and loss of follow-up^16,21,27^. It was reported that detecting the diverse rates of congenital anomalies in isotretinoin exposure between previous studies before 2006 and current studies might arise from the differences in isotretinoin daily dose and exposure week^18^. Our study revealed that there is no relationship between isotretinoin exposure and daily dose, cumulative dose, and exposure time.

With the advancement of pregnancy prevention programs, nowadays, it is possible to detect isotretinoin exposure during pregnancy early in the pregnancy. As a result, fetuses having a potential congenital abnormality are terminated in the early part of the pregnancy. It is also known that termination rates are higher in risky period exposure ^22^. Therefore, congenital anomaly rates in isotretinoin exposure should not be evaluated independently from termination rates. A meta-analysis indicated that isotretinoin-exposed pregnancy termination rates were 44-82% ^15^. In our study, the termination rate in the isotretinoin exposure before the pregnancy group was 9.3%, in the isotretinoin exposure during pregnancy was 32.5%, and in the control group was 2.7%. Although there is a statistically significant difference detected in both the isotretinoin exposure during the pregnancy group and the isotretinoin exposure before the pregnancy group compared to controls, our termination rate was one of the lowest termination rates in studies so far. The low termination rates and no missing values in our study result in more accurate congenital anomaly detection by reducing bias.

During teratological counseling, it is explained that there is an increased risk of congenital anomalies resulting from isotretinoin exposure (28-35%), and this risk is high and there is an indication for termination. However, some families decided to continue their pregnancies and give birth to babies with congenital anomalies. For this decision, first-contact physicians impact, wrong interpretation of religious information, close relatives and neighborhood pressure, lack of information, and socioeconomic status rather than reliance or mistrust of a counselor might play a role^6,28,29^.

During the first examination, the recommendation for termination by first-contact physicians to pregnant women who stopped isotretinoin a month or more before their conceptions is another significant topic that should be discussed for community health issues. Many pregnant women had fear and worries about their pregnancies after the termination recommendations. However, after the teratological counseling, most of the women decided to continue their pregnancies. Termination recommendations in women with isotretinoin exposure before their pregnancies were 60%, and 15.66% of them terminated their pregnancies without any indications. Surprisingly, termination recommendations for women with isotretinoin exposure before the pregnancy were not only higher than for the control group but also higher than for women with isotretinoin exposure during their pregnancies. This might be associated with the recommendation of terminations implemented in earlier periods of the pregnancies. These pregnant women who applied to our center for teratological consultation were informed about the duration of the drug’s stay in the tissues and its teratogenic effect potential, and it was stated that an increase in anomaly risk was not expected, therefore there was no termination indication. More than 90% of women for whom termination was recommended decided to continue their pregnancies after the teratological counseling. After the birth, no specific features were recorded unique to RA embryopathies in babies born from these pregnancies. This result indicated that teratological counseling is a beneficial community service for inhibiting fears and worries resulting from a lack of information and pregnancy losses^28^.

In the teratological consultation given to pregnant women using isotretinoin during pregnancy, it was stated that the risk of anomaly is 20-35%, the risk of neurocognitive disorder is 30%^23^, and the risk of miscarriage up to 20%^22^ if the drug is used in the critical period. The critical period for isotretinoin is a time range from the second week to the fifth week after conception^30^, and therefore it was also explained to pregnant woman that before the critical period, adverse pregnancy outcome risks were lower than that but higher than community risk. In our study, it was observed that while isotretinoin exposure during the pregnancy increased the congenital anomaly rates compared to control group, isotretinoin exposure before the pregnancy did not increase miscarriage risk. Our study also indicated that isotretinoin exposure during the pregnancy led to increased rates of miscarriages compared to the control group; however, when isotretinoin exposure before the pregnancy and the control group were compared, there was no difference found between the groups. However, isotretinoin exposure before the pregnancy or during the pregnancy both increased the risk of prematurity compared to control group, which one previous study found to be 7%^16^.

Although there have been many novel phenotypical features reported in RA embryopathy, the reasons for these features were not investigated in previous studies. However, these novel features of RA embryopathy might stem from retinoic acid exposure, genetic variation, or incidentally. Therefore, it is essential to differentiate those possible causes. Genetic alterations cause some types of syndromes, and agents affect RA signaling pathways to mimic the clinical characteristics of RA embryopathy^31^. Therefore, experts recommend that genetic cytogenetic and molecular tests and segregation analysis of families together be implemented to exclude genetic etiologies^32,33^. In our study, cytogenetic and molecular tests were performed together in five cases with new phenotypic findings, and it was shown that the new phenotypic finding in three cases born to mothers who used isotretinoin during pregnancy may be associated with a genetic variant[Table 3].

There are many strengths in our study, including study design (retrospective cohort), long-term follow-ups, implementation of discrimination between isotretinoin exposure time (before the pregnancy and during the pregnancy), observation of one of the lowest termination rates, and finding anomalies in babies by evaluating both clinical and laboratory methods (molecular and cytogenetic methods). However, there are some limitations in our study, which include termination recommendations based on expectant mothers’ verbal statements, some pregnancies were terminated, and some of the pregnancy losses occurred before implementing genetic analysis.

As a conclusion of the study, study design, teratological counseling, and advanced molecular techniques are beneficial for detecting the relationship between congenital anomalies and isotretinoin exposure. Our data also supports that there is not scientific proof of termination suggestions without discriminating the isotretinoin exposure period (before the pregnancy or during the pregnancy). Therefore, teratological counseling is a cheap, practical, and effective community health service for reducing congenital anomalies and unwanted pregnancy losses.

## Data Availability

All data produced in the present study are available upon reasonable request to the authors

## Contributor statement

MTA, and MS conceived and designed the analysis. MTA collected data and performed the statistical analysis. MTA wrote the paper. MTA, AK, MS drafted, revised, and approved the final version.

## Acknowledgements

Our kind respect for all efforts on preventing isotretinoin exposure in pregnancy for all dermatologists, and our special thanks to our pregnant women included in the study.

